# Integrated Genomic Analyses Identify High-Risk Factors and Actionable Targets in T-Cell Acute Lymphoblastic Leukemia

**DOI:** 10.1101/2021.07.17.21260159

**Authors:** Haichuan Zhu, Bingjie Dong, Yingchi Zhang, Mei Wang, Jianan Rao, Bowen Cui, Yu Liu, Qian Jiang, Weitao Wang, Lu Yang, Anqi Yu, Zongru Li, Chao Liu, Leping Zhang, Xiaojun Huang, Xiaofan Zhu, Hong Wu

**Affiliations:** The MOE Key Laboratory of Cell Proliferation and Differentiation, School of Life Sciences, Peking University, Beijing 100871, China; Peking-Tsinghua Center for Life Sciences, Peking University, Beijing 100871, China; State Key Laboratory of Experimental Hematology, Institute of Hematology and Blood Diseases Hospital, Chinese Academy of Medical Sciences & Peking Union Medical College, Tianjin 300020, China; Pediatric Translational Medicine Institute, Shanghai Children’s Medical Center, School of Medicine, Shanghai Jiao Tong University, Peking University People’s Hospital, Beijing 100044, China; Shanghai Key Laboratory of Clinical Molecular Diagnostics for Pediatrics, Peking University People’s Hospital, Beijing 100044, China; Peking University Institute of Hematology, National Clinical Research Center for Hematologic Disease, Peking University People’s Hospital, Beijing 100044, China; Department of Pediatrics, Peking University People’s Hospital, Beijing 100044, China

## Abstract

T cell acute lymphoblastic leukemia (T-ALL) is an aggressive hematologic malignancy often associated with poor outcomes. To identify high-risk factors and potential actionable targets for T-ALL, we perform integrated genomic and transcriptomic analyses on samples from 165 Chinese pediatric and adult T-ALL patients, of whom 85% have outcome information. The genomic mutation landscape of this Chinese cohort is very similar to the Western cohort published previously, except that the rate of *NOTCH1* mutations is significant lower in the Chinese T-ALL patients. Among 47 recurrently mutated genes in 7 functional categories, we identify *RAS* pathway and *PTEN* mutations as poor survival factors for non-TAL and TAL subtypes, respectively. Mutations in the *PI3K* pathway are mutually exclusive with mutations in the *RAS* and *NOTCH1* pathways as well as transcription factors. Further analysis demonstrates that approximately 43% of the high-risk patients harbor at least one potential actionable alteration identified in this study, and T-ALLs with *RAS* pathway mutations are hypersensitive to MEKi *in vitro* and *in vivo*. Thus, our integrated genomic analyses not only systematically identify high-risk factors but suggest that these high-risk factors are promising targets for T-ALL therapies.

## Introduction

T-cell acute lymphoblastic leukemia (T-ALL) is a heterogeneous disease caused by accumulated genetic alterations in T progenitor cells[1, 2]. The incidence of T-ALL is approximately 10%-15% in pediatric ALL and 25% in adult ALL[3, 4]. Despite advances in T-ALL treatment, approximately 20% of pediatric and 40% of adult patients are expected to have poor prognosis[5, 6].

Recent unbiased large-scale genomic landscape studies, some in combination with transcriptome analyses, have revealed frequently mutated genes and dysregulated pathways associated with several major subtypes of T-ALLs[7-9]. Other prognosis-based sequencing analyses are mainly focused on adult T-ALL. Mutations in the *JAK/STAT, RAS/PTEN, TP53, IDH2* and *DNMT3A* genes are correlated with worse survival, while mutations in the *NOTCH* pathway as well as *TCR* gamma and *CDKN2A/CDKN2B* homozygous deletion are related with better survival in adult T-ALL patients[10-12]. Chromosomal rearrangements are another frequent genetic alterations found in T-ALL, which lead to dysregulated transcription factors[1]. Dysregulated *HOXA* cluster expressions, either by fusion events or 3D genome alterations, are associated with poor survival of T-ALL[13]. Additionally, *MLL* related rearrangement and *SPI1* fusions were related with worse survival[9, 14]. ETP ALL, a subtype of T-ALL characterized by the lack of mature T cell markers and the expression of stem and myeloid-lineage genes, has also been reported to have poor prognosis[15, 16]. However, systematic investigation of the genetic landscape underlying poor prognosis of T-ALL, especially pediatric T-ALL, is needed for molecular-based prognosis and designing new targeted therapeutic strategies.

In this study, we conducted integrated whole-exome sequencing (WES) and RNA sequencing (RNA-seq) analyses on a large Chinese cohort to identify those high-risk factors associated with poor prognosis of T-ALL. As many of the high-risk factors identified are potential drug targets, our study may shed light on the molecular-based prognosis of T-ALL, which would enable the design of new therapeutic strategies.

## Materials and methods

### T-ALL patient samples

Primary and remission samples used for this study were obtained from the Institute of Hematology at Peking University (n=66) and the Tianjin Institute of Hematology (n=99) from 2010 to 2018. Among 165 patient samples, 33 were from adult (age >= 18) and 128 were from pediatric (age < 18) T-ALL patients, four patients missed age information. The study protocols were approved by the ethics committees of these two institutions. All patients gave written informed consent for treatments and sample collections.

The diagnosis of T-ALL was made by morphological, immunophenotypical and cytogenetic analyses of bone marrow specimens according to the WHO classification[17]. ETP ALL and non-ETP ALL status were determined according to a previous publication[15]. Immunophenotypes were evaluated by 8-color multi-parameter flow cytometry analysis. Other detailed patient-related information can be found in Table 1 and supplemental Table 1.

**Table 1.**
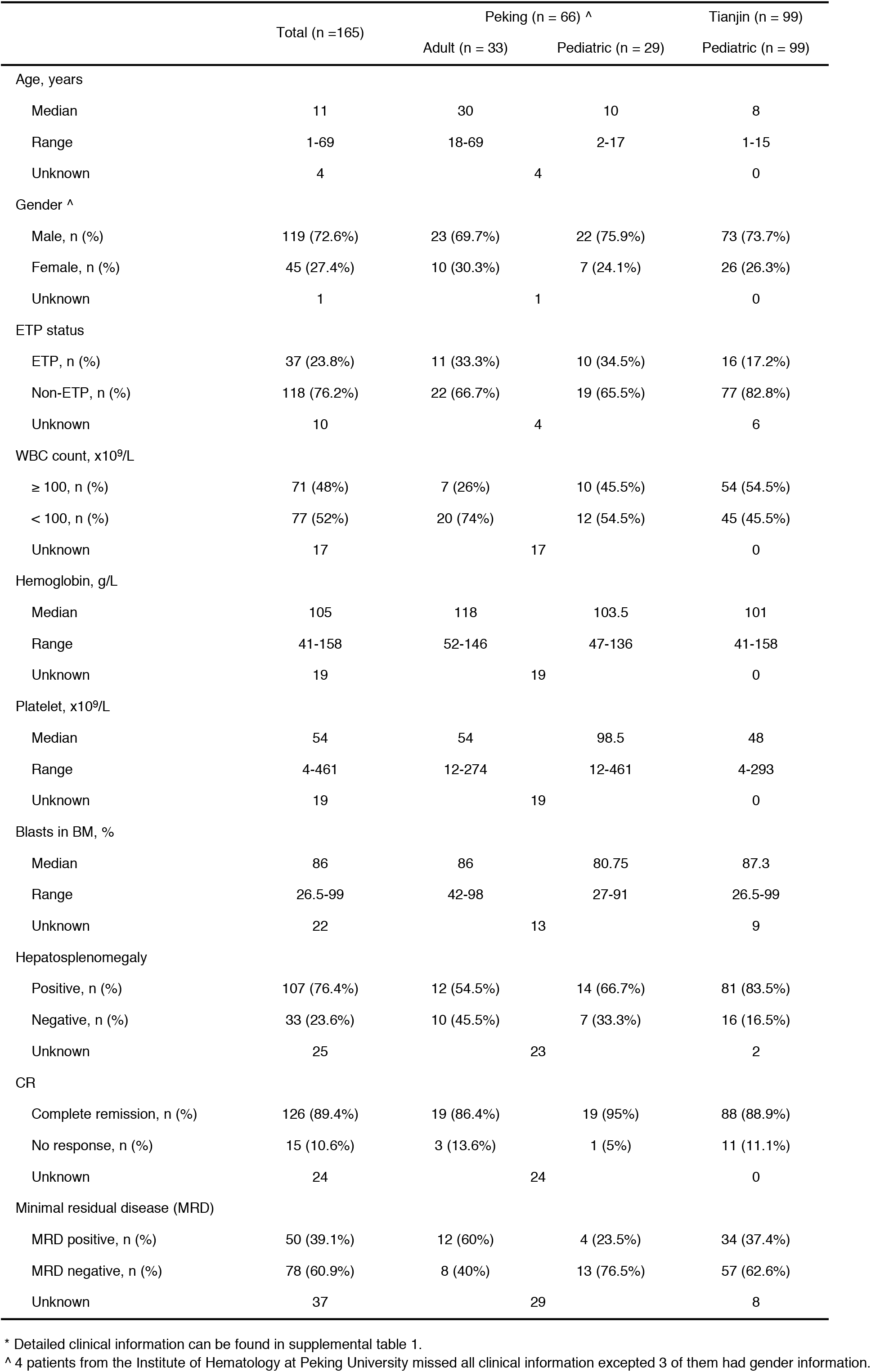
Clinical Characteristics of the T-ALL Patient Cohort *

### Treatment

The treatment protocol included induction and post-remission therapy. For pediatric patients, the Chinese Children Leukemia Group CCLG-2008 protocol was used for 50 patients between April 2008 and December 2014, and the CCLG-2015 protocol was used for an additional 49 patients after 2015. Details of the CCLG-2008 and CCLG-2015 treatment regimens have been published previously[18, 19].

For adult patients, CODPL regimens (cyclophosphamide, daunorubicin, vincristine, prednisone and L-asparaginase) were used as the induction therapy. After remission, patients received consolidation therapy, including a modified hyper-CVAD regimen (fractionated cyclophosphamide, vincristine, doxorubicin, and dexamethasone alternating with high-dose methotrexate and cytarabine for four cycles). After consolidation chemotherapy, patients received a maintenance regimen (vindesine, prednisone, mercaptopurine, and methotrexate) for 2 years. Based on physician recommendations as well as patients’ desires, some patients with available donors received allo-HSCT after at least two cycles of consolidation therapy according to the protocol published previously[20].

### Whole-exome sequencing (WES)

Genomic DNA was extracted using a DNeasy Blood & Tissue Kit (69504, Qiagen). The quality of the DNA samples was measured with agarose gel electrophoresis. One microgram of high-quality DNA from each sample was used for exome enrichment and library preparation following the manufacturer’s protocols of SureSelectXT. Human All Exon V6 (Agilent Technologies, 5190-8865) and NEBNext® Ultra™ DNA Library Prep Kit (NEB, E7645) were used for the library construction. The WES library was then sequenced on the HiSeq X Ten platform after performing quality control analysis.

### Whole-exome sequencing data analysis

Paired-end reads were aligned with BWA v0.7.15[21] to the human reference genome hg19. Duplicates were marked with Picard tool v2.9.2. GATK toolkit v3.7.0[22] was used for realignment and base recalibration. Single-nucleotide variants (SNVs) were identified by MuTect v1.1.7[23] and Bambino v1.06[24] using 42 primary-remission paired samples. Indels were identified by Strelka v1.0.15[25] and Bambino v1.06[24]. Significantly mutated genes were identified by MuSiC[26].

For unpaired samples, we created a panel of normal controls using the 42 remission samples. We then used the MuTect2 algorithm to identify SNVs and indels. Variants were filtered by removing those that met one of the following conditions: (1) < 6 supporting reads in tumor samples; (2) < 20 depth in tumor samples; (3) > 2 supporting variant reads in normal samples; (4) < 10 depth in normal samples; (5) identification as variants in the 1000 Genome East Asian Project (August 2015 release) and not in the COSMIC database (version 70); and (6) identification as variants in the ExAC nonTCGA East Asian database (version 0.3) and not in the COSMIC database. We used Medal Ceremony[27] to annotate driver sites. All the variants were verified by the Integrative Genomics Viewer[28]. We used GATK4 to identify copy number variations, recurrent copy number variations were calculated by GISTIC2[29].

### mRNA sequencing

RNA-seq library was established as previously described[30]. In brief, total RNA was isolated from each sample using a RNeasy Mini Kit (Qiagen, 74104). Oligo(dT)25 cellulose beads were used for the isolation of mRNA from 1 µg of total RNA. After cDNA generation, RNA-seq library was prepared using an NEBNext® Ultra™ DNA Library Prep Kit for Illumina (NEB, E7645) following the manufacturer’s protocol, and was sequenced on a HiSeq X Ten platform.

### T-ALL PDX models and in vivo treatment

To establish PDX mouse models, 6-to 8-week-old NOD/SCID/IL-2Rγ-null (NCG) mice (Jiangsu GemPharmatech Co, Ltd, China) were sub-lethally irradiated (2 Gry; once one day before transplantation), and 5×10^5^ T-ALL cells harvested from patients’ bone marrow were intravenously injected. Ten days after the initial transplantation, mice were randomly assigned into control and treatment groups. PD0325901 (25mg/kg) was administrated once daily. Leukemia development was monitored daily by physical appearance, and weekly by peripheral blood smear and FACS analysis using anti-human CD7 antibody. T-ALL was confirmed when leukemia burden reached >20% in peripheral blood, and the spleen was harvested for analysis as described previously[30]. The experiments were performed with the approval of the Animal Ethics Committee of Peking University under the protocol of ID LSC-WuH-1.

### mRNA sequencing data analysis

RNA sequencing reads were aligned to human hg19 with MapSplice v2.1.8[31]. Gene expression was quantified by RSEM[32] using the transcript model TCGA GAF (https://gdc.cancer.gov/about-data/data-harmonization-and-generation/gdc-reference-files). Gene fusion was detected by CICERO[33]. Differential expression analysis was performed using R package DESeq2 v1.22.1[34]. KEGG enrichment analysis was performed by clusterProfiler[35].

MuTect2 and RNAIndel[36] were applied for SNV and Indel analysis and annotated with VEP[37]. Variants were further filtered by removing those that met one of the following conditions: (1) in Ig/TCR region; (2) < 3 supporting variant reads; (3) < 8 reads depth; (4) > 0.001 allele frequency in 1000 Genome; (5) not in coding or splicing regions; (6) matching variant in the internal artifact blacklist representing mutation artifact. We then selected variants meets any of the flowing conditions: (1) vaf greater than 0.1, predicted as ‘deleterious’ and ‘possibly damaging’ in SIFT and PolyPhen respectively, in the internal tumor associated gene list; (2) recurrent (n>=3) in the COSMIC[38] database or in previously published datasets including PCGP[39] or TARGET[40].

We used fusion events and the expression of transcription factors to classify patients into 4 subtypes: (1) LMO2/LYL1: patients with both *LMO2* and *LYL1* upregulation; (2) HOXA: patients with *HOXA* activation related fusion events or *HOXA* gene upregulation; (3) LTX: patients with *TLX3* activation related fusion events, or patients with *TLX1, TLX3* or *NKX2-1* upregulation; (4) TAL: patients with *TAL1* or *TAL2* activation related fusion events, or patients with T*AL1* or *TAL2* upregulation[8, 41].

### Statistical and survival analyses

Fisher’s exact test was used to identify associations between mutations or pathway alterations with age, MRD status and immunophenotype. MRD positivity status as evaluated at induction therapy about one month, MRD positive was defined with MRD larger than 0.01%.

Survival analysis was performed using a Cox regression model and presented as overall or event-free survival as outcomes. Overall survival was defined as the time from diagnosis to death from any cause. Event-free survival was defined as the time from diagnosis to treatment failure, relapse, or death from any cause. Variables tested in the multivariable Cox regression model included risk factors, age (pediatric vs. adult), gender, white blood cell count (WBC< 100×10^9^/L)[42], hemoglobin (<100g/L), platelet count (<100×10^9^/L)[16] and hepatosplenomegaly. Patients who received transplantation were excluded from survival analysis.

### Data access

The raw sequence data reported in this paper have been deposited in the Genome Sequence Archive [43]in National Genomics Data Center[44], China National Center for Bioinformation / Beijing Institute of Genomics, Chinese Academy of Sciences, under accession number HRA000122 that are publicly accessible at https://bigd.big.ac.cn/gsa.

## Results

### Clinical characteristics of T-ALL patients

To identify high-risk factors in T-ALL, we analyzed 165 pediatric and adult T-ALL samples collected from two Chinese hospitals between 2010 and 2018. There were 33 adult and 128 pediatric patients, with 4 patients missing information. Among 128 pediatric patients, 99 patients from Tianjin were participants of the CCLG-2008 and CCLG-2015 clinical trials[18, 19]. The overall complete remission rate in the entire cohort was 89.4%, MRD negative rate was 60.9% after first round of the treatment and ETP rate was 23.8%. The detailed clinical information could be found in Table 1 and Supplemental Table 1.

### Genomic landscape of Chinese T-ALL

To investigate the genomic landscape-associated with different outcomes, we conducted whole-exome sequencing (WES) and RNA-seq analyses, and the overall experimental design was illustrated in supplemental Figure 1A. WES analysis on 42 paired samples identified 585 somatic coding mutations, including 471 nonsynonymous single-nucleotide variants (SNVs) and 114 insertions/deletions (indels) (Supplemental Table 2). For the additional 79 samples without matched remission controls, we also conducted WES analysis, using pooled 42 remission samples as germline and background controls, and performed mutation calling. In total, we identified 47 recurrently mutated genes (Supplemental Table 3), which belonged to seven functional categories (Figure 1A).

**Figure 1.**
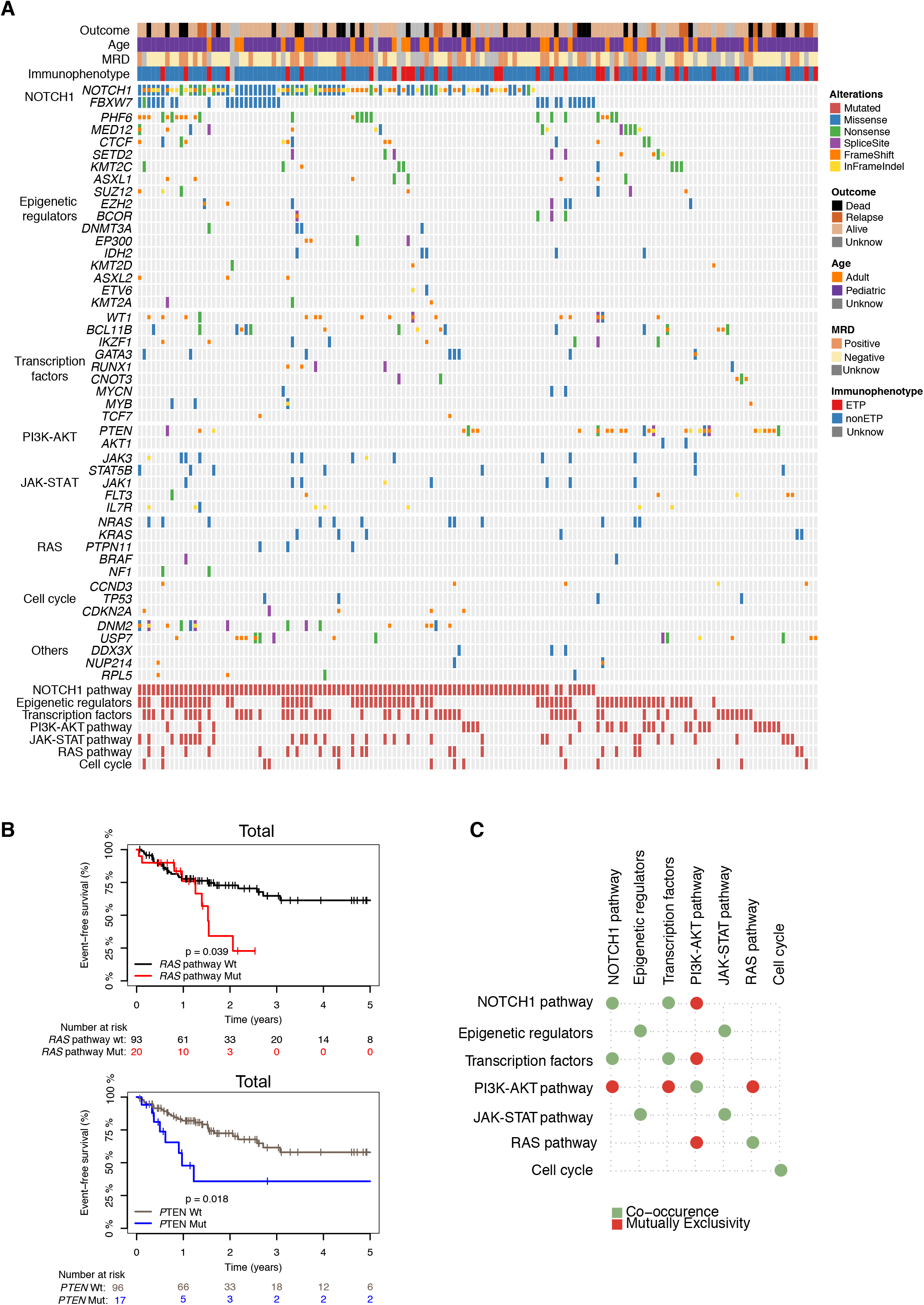
Mutational landscape-associated in T-ALL. (A) Recurrently mutated genes in T-ALL were ordered by functional categories shown on the left. Synonymous mutations were excluded. Lower part summarized mutations in each functional category. (B) Kaplan-Meier event-free survival of T-ALLs with (red) or without (black) *RAS* pathway mutations (upper) and with (blue) or without (grey) *PTEN* mutations (lower) in the entire cohort. (C) Co-occurring (green) or mutually exclusive (red) pathway alterations (p<0.05; two-sided Fisher’s exact test).

The overall somatic mutation burden and signature in this Chinese T-ALL cohort were very similar to the previous report[40] (Supplemental Figure 1B-C). However, the mutation rate of *NOTCH1* was lower, while the mutation rates of *SETD2, CDKN2A, ASXL1* and *TP53* were higher in the Chinese cohort compared to the White population (Supplemental Figure 1D)[7, 8]. WES-based CNV analysis also identified recurrent deletions in the *CDKN2A*/*CDKN2B, TCRA, TCF7* and *SUZ12* loci (Supplemental Figure 1E), consistent with a previous publication [8]. High frequent chromosome 14 and 15 amplifications were also observed, but the biological significance of this finding requires further investigation. For the 124 samples with RNA-seq data, we followed the steps described in the Method section to identify the potential driver mutations. Mutations identified in this way were largely consistent with those identified by WES analysis (Supplemental Figure 1F).

### Recurrent alterations-associated with clinical outcomes of entire population

Among the seven functional categories (Supplemental Table 3), *NOTCH* pathway mutations had the highest mutation frequency, affecting more than half of the patients, followed by mutations in the epigenetic regulators and transcription factors. Mutations in the *PI3K, JAK-STAT* and *RAS* pathways were presence in 17%, 20% and 14% of the T-ALL samples, respectively. We observed that *PTEN* mutation was the dominant factor affecting the *PI3K* pathway. We also identified that 8% patients harbored cell cycle related mutations (Supplemental Table 3).

We then investigated the relationship between the recurrent pathway mutations and clinical outcomes in the entire population (n=113), and found that *RAS* pathway and *PTEN* alterations were related with event-free survival (EFS) (Figure 1B, Supplemental Figure 1G and Supplemental Table 5 and 6). Interestingly, mutations in the *PI3K* pathway were mutually exclusive from mutations in the *RAS* and *NOTCH* pathways as well as transcription factors (Figure 1C).

Sub-clonal mutations are defined by mutant allele fraction (MAF) of less than 0.3[8, 45]. Interestingly, most *RAS* pathway mutations were monoclonal mutations, except two patients harboring *NRAS* and *NF1* mutations, and 5/23 samples were sub-clonal. On the other hand, nearly half samples with *PTEN* mutations carried two or more than two distinct *PTEN* mutations and 4/27 samples were sub-clonal (Supplemental Figure 1I).

At individual gene level, we found that patients with *JAK3* (n=7), *ASXL1* (n=7) mutations might be related with worse outcome (Supplemental Table 5 and 6).

### Mutations associated with age and MRD

Age is known to associate with worse outcome in T-ALL[46]. In our cohort, 82% adult T-ALL were dead or relapsed within 3 years and had poor outcome compared with pediatric T-ALL (Figure 2A; Supplemental Figure 2A). We found a higher mutation rates of epigenetic regulators in adult T-ALL, e.g., *DNMT3A* and *IDH2* mutations were only detected in adult T-ALL while *MED12* mutations were highly enriched in adult T-ALL (Figure 2B and Supplemental Figure 2B). In addition, we also found higher mutation rates of *JAK3* and *JAK-STAT* pathway in adult T-ALL (Figure 2B and Supplemental Figure 2B).

**Figure 2.**
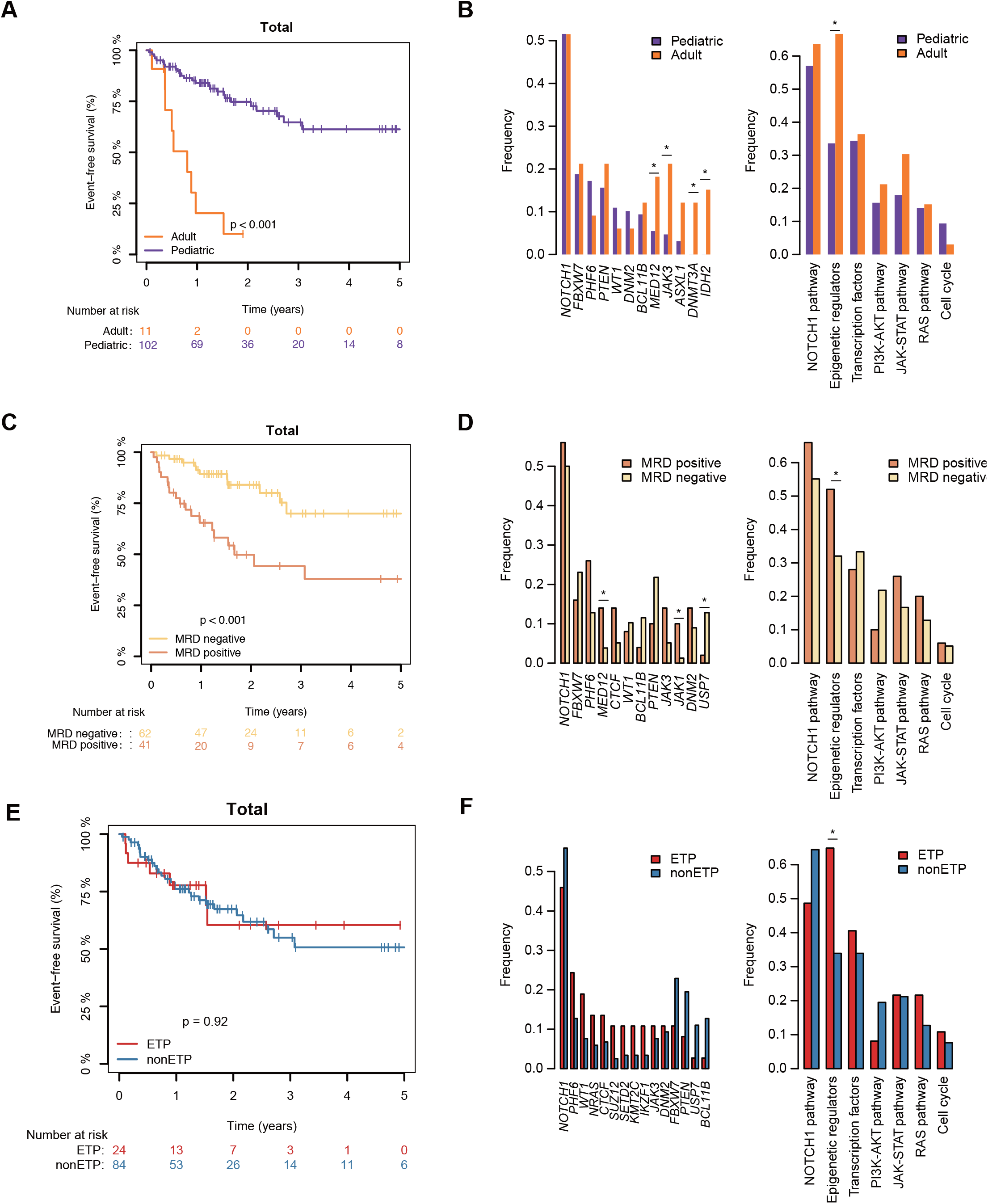
Relationship between recurrent mutations and clinical features. (A) Kaplan-Meier event-free survival curves of adult (orange) and pediatric T-ALLs (purple). (B) Bar graph showed the different rates of gene mutation (left) or mutation-associated functional category (right) in adult (orange) and pediatric (purple) T-ALLs. (C) Kaplan-Meier event-free survival curves of MRD positive (brown) and MRD negative (yellow) T-ALLs. (D) Bar graph showed the different rates of gene mutation (left) or mutation-associated functional category (right) in MRD positive (brown) and MRD negative (yellow) T-ALLs. (E) Kaplan-Meier event-free survival curves of ETP (red) and non-ETP ALLs (blue). (F) Bar graph showed the different rates of gene mutation (left) or mutation-associated functional category (right) in ETP (red) and non-ETP ALLs (blue). *, p<0.05

As for pediatric T-ALL, *RAS* pathway mutations were associated with both overall survival (OS) and EFS in all pediatric T-ALL (n=102) and those pediatric patients who participated in the clinical trials (n=91), while *PTEN* mutations were only associated with EFS of pediatric patients participated in the clinical trials (Supplemental Figure 1H; Supplemental Table 5 and 6).

MRD is a clinical feature routinely used in monitoring the T-ALL patients[47], and we found that the MRD positive rate was much higher in adult T-ALL than that of pediatric T-ALL (60% vs. 35%; Table 1). MRD positivity was related with worse outcome in the entire cohort and pediatric T-ALL (Figure 2C and Supplemental Figure 2C-D). *MED12* and *JAK1* mutations were positively correlated with MRD positivity, while *USP7* mutation was negatively correlated with MRD positivity (Figure 2D and Supplemental Figure 2E).

ETP ALL has been reported as an aggressive subtype of T-ALL with poor prognosis[15, 16, 48], although another study found no difference in the overall survival between ETP ALL and non-ETP ALL[49]. Despite the higher incidences of ETP ALL in adult T-ALL than that in pediatric T-ALL, 33.3% and 21%, respectively, and enriched epigenetic regulators mutations, we did not find significant differences in the outcomes between ETP ALL and non-ETP ALL both in the entire cohort and in pediatric T-ALLs (Table 1; Figure 2E-F and Supplemental Figure 2F-G). Thus, ETP is not a significant contributing factor for poor prognosis in our cohort.

### Recurrent mutations-associated with major T-ALL subtypes

Previous study demonstrated that chromosome translocation-mediated gene fusions were major drivers of leukemogenesis[1]. According to the fusion events-associated dysregulated transcription factors, T-ALLs could be separated into six major subtypes, i.e., LOM2/LYL1, HOXA, TLX3, TLX1, NKX2-1 and TAL1 subtypes [8, 41]. Utilizing the RNA-seq data from 124 patients, we identified 156 fusion events in 86 (69.4%) samples (Figure 3A-B and Supplemental Table 7). Most of these recurrent fusion events were associated with HOXA and TAL1 subtypes, while only one *TLX3* related fusion event was detected in 21 cases with *TLX3* upregulation, which was much lower than the previous report[8] (Figure 3B, top listed fusion events identified in this study). We also detected *TCF7-SPI1* translocation in 3 cases (2.4%), which was reported as a high risk factor in a previous publication[9] (Figure 3A).

**Figure 3.**
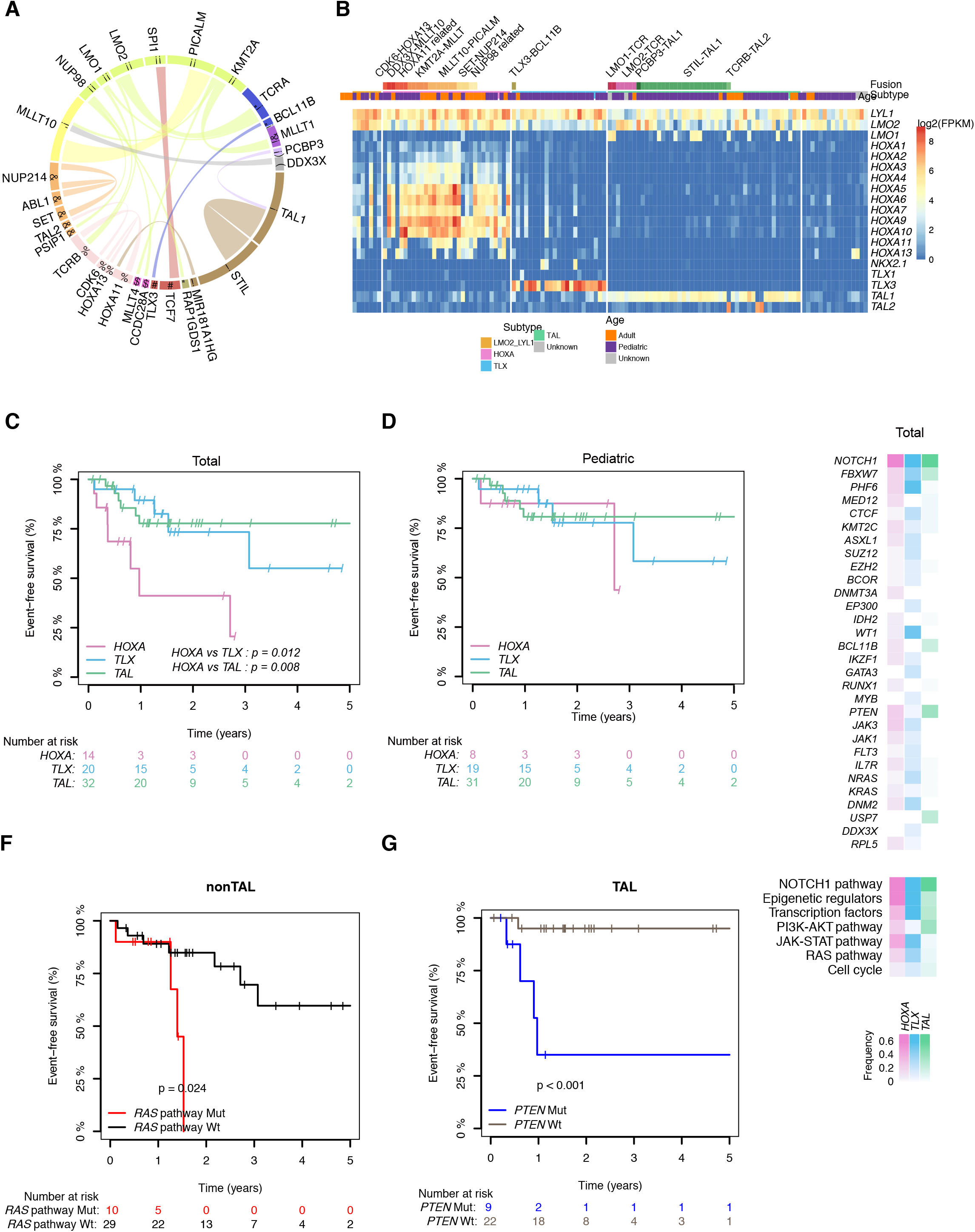
The association of recurrent mutations and major subtypes of T-ALL. (A) Circos plot of the oncogenic fusion events discovered by RNA-seq, ordered by chromosome. Ribbon widths were proportional to the frequency of each fusion event. (B) Heatmap showed the major fusion events and associated dysregulated transcription factors expression, annotated by subtypes and age. (C) and (D) Kaplan-Meier event-free survival curve of entire cohort (C) or pediatric T-ALLs (D) with HOXA (pink), TLX (blue) and TAL subtypes (green). (E) Heatmap showed the rates of gene mutation (top) or mutation-associated functional category (bottom) in HOXA, TLX and TAL subtypes. (F) Kaplan-Meier event-free survival curve of pediatric T-ALLs with (red) or without (black) *RAS* pathway mutations in non-TAL subtype. (G) Kaplan-Meier event-free survival curve of pediatric T-ALLs with (blue) or without (grey) *PTEN* mutations in the TAL subtype.

Since there were only 7 samples in the LMO2/LYL1 subtype, we analyzed EFS of patients within the HOXA (n = 14), TLX (n = 20) and TAL subtypes (n = 32). For the entire cohort, the HOXA subtype showed a worse EFS when compared to the TLX and TAL subtypes (Figure 3C), and KEGG analysis showed that HOXA subtype was enriched for term such as transcriptional misregulation in cancer and MAPK signaling pathway (Supplemental Figure 3A-B). However, the survival difference was diminished when only pediatric T-ALL cases were analyzed (Figure 3D and Supplemental Figure 3C).

We also studied mutations that were preferentially associated with each subtype. We found higher mutation frequencies of *IDH2, DNMT3A* and *FLT3* in the LMO2/LYL1 subtype (Supplemental Figure 3E), *MED12* and *RPL5* mutation in the HOXA subtype, *PHF6, CTCF, EP300, WT1, GATA3*, and *NRAS* mutation in the TLX subtype, while *USP7* mutation only in the TAL subtype (Figure 3E, upper). There were also higher number of adult T-ALL found in the HOXA subtype with 15/31 adult in HOXA, while only 2/23 and 6/45 adult T-ALL were found in TLX and TAL subtypes, respectively (Figure 3B and Supplemental Figure 3D).

Interestingly, *RAS* pathway alterations were associated with poor survival of non-TAL subtypes while *JAK3* mutation was associated with poor prognosis of TLX subtype (Figure 3F and Supplemental Figure 3F). The TAL subtype showed the highest 5-year event free survival rate (Figure 3C), but the high-risk cases within this subtype were all associated with *PTEN* mutations (Figure 3G), and were mutually exclusive from *USP7* or *BCL11B* mutations (Supplementary Figure 3G). Comparing with patients with *PTEN* mutation in the TAL subtype, most patients with *USP7* or *BCL11B* mutations were MRD negative (Figure 2D) and showed better outcome (Supplementary Figure 3H).

### T-ALL with RAS pathway mutations are hypersensitive to MEK inhibition

T-ALLs with the *PI3K* pathway activation are sensitive to those PI3K, AKT and mTOR inhibitors both *in vitro* and *in vivo[30, 50, 51]*. To determine whether *RAS* pathway mutations could be potential drug targets, we investigated the nature of the mutations in our cohort and found most of them were gain-of-function mutations (Figure 4A and Supplemental Figure 4A). Many of the same mutations could also be found in human T-ALL lines (Figure 4A). Importantly, T-ALL cell lines with *RAS* pathway mutations, such as DND-41, MOLT-3, MOLT-4, MOLT-13, KE-37, CCRF-CEM, and P12-ICHIKAWA, were more sensitive to MEK/ERK inhibitors CI-1040, PD0325901, Refametinib and Trametin with lower IC50 (Figure 4B; Supplementary Table 10)[52]. PF-382 was an exception as it was resistant to CI-1040 and PD0325901 but relatively sensitive to Refametinib and Trametin(Figure 4B; Supplementary Table 10).

**Figure 4.**
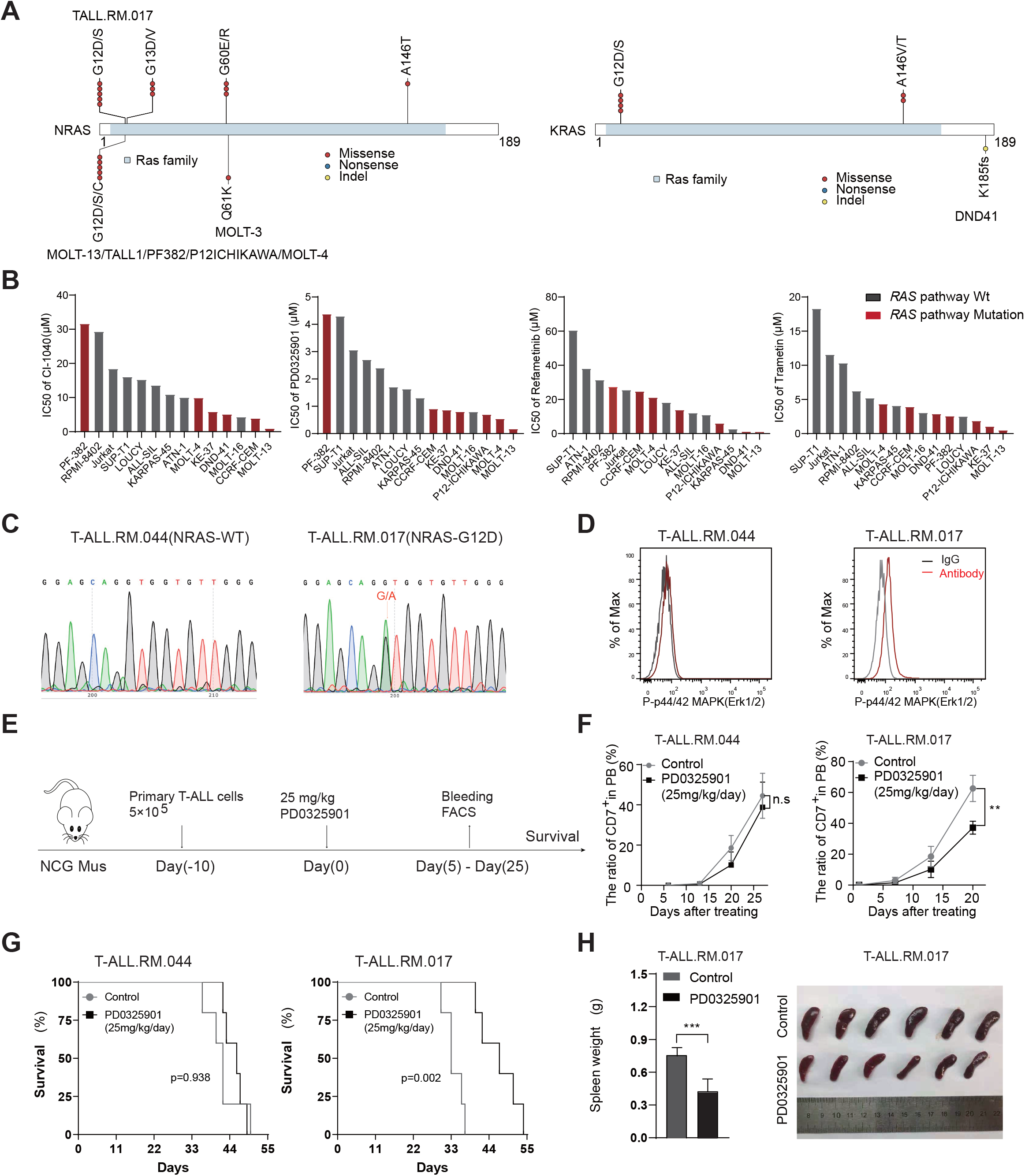
T-ALLs with mutations in the RAS pathway are sensitive to MEK inhibition. (A) Mutation profile for *NRAS* (left) and *KRAS* (right) in T-ALL samples (top) and T-ALL cell lines (bottom; CCLE database [https://portals.broadinstitute.org/ccle]). (B) The IC50 values of 4 MEK inhibitors in 14 T-ALL cell lines. Red, cell line with *RAS* pathway mutation; grey, WT for *RAS* pathway. (C) Representative DNA sequencing chromatograms of *N-RAS* WT (T-ALL.RM.044) and mutant (T-ALL.RM.017) samples, showing a mono-allelic G12D mutation. (D) Intracellular FACS analyses of P-ERK levels in the *NRAS* WT (T-ALL.RM.044) and mutant (T-ALL.RM.017) patient samples. Gray line: isotype control. (E) A schematic outline of *in vivo* drug treatment using the PDX models. n=6 per group. (F) The proportion of human CD7^+^ leukemic blasts in the peripheral blood were measured by FACS from day 0 to day 25 in control and drug treatment cohorts. **, p<0.01. (G) Kaplan-Meier survival curves of PDX models treated with placebo (control) and PD0325901. (H) Spleens from control and treatment groups were weighed and photographed. ***, p< 0.001.

In our cohort, 9 out of 18 patients carried *N-RAS/K-RAS* mutations at amino acid Gly12, a hot spot mutation known to associate with RAS pathway activation. We further confirmed mono-allelic mutation by Sanger sequencing, and demonstrated the RAS/MAPK pathway activation by intra-cellular FACS analysis of T-ALL-RM-017 (RAS mutated) and T-ALL-RM-044 (RAS WT) samples (Figure 4C-D). Using PDX mouse models generated from these T-ALL bone marrow samples, we found that T-ALL-RM-017 was more sensitivity to the MAPK pathway inhibitor PD0325901 than that of T-ALL-RM-044 *in vivo* (Figure 4E-F). PD0325901 could also significantly prolong the survival of T-ALL-RM-017 leukemia mice (Figure 4G) by reducing leukemic burden, as evidenced by reduced spleen weight and size in the T-ALL-RM-017 PDX mice model (Figure 4H). Taken together, these results suggest that T-ALLs with *RAS* pathway mutations are hypersensitive to RAS/RAF/MAPK pathway inhibition.

## Discussion

Taken the advantage of available survival related information in this Chinese cohort, we were able to identify those high risk factors that significantly contributed to poor prognosis of T-ALL (Figure 5). Survival analysis demonstrated that adult T-ALL had median OS and EFS time less than 1 year, while MRD-positive patients had the median OS and EFS time around 2 years in the entire population and 2 – 3 years in the pediatric patients (Figure 5A). *MED12* and *JAK1/JAK3* mutations were significantly enriched in adult and MRD-positive patients, while *DMNT3A* and *IDH2* mutations were only present in adult T-ALL patients in our cohort (Figure 2B and Supplemental Figure 2B and E).

**Figure 5.**
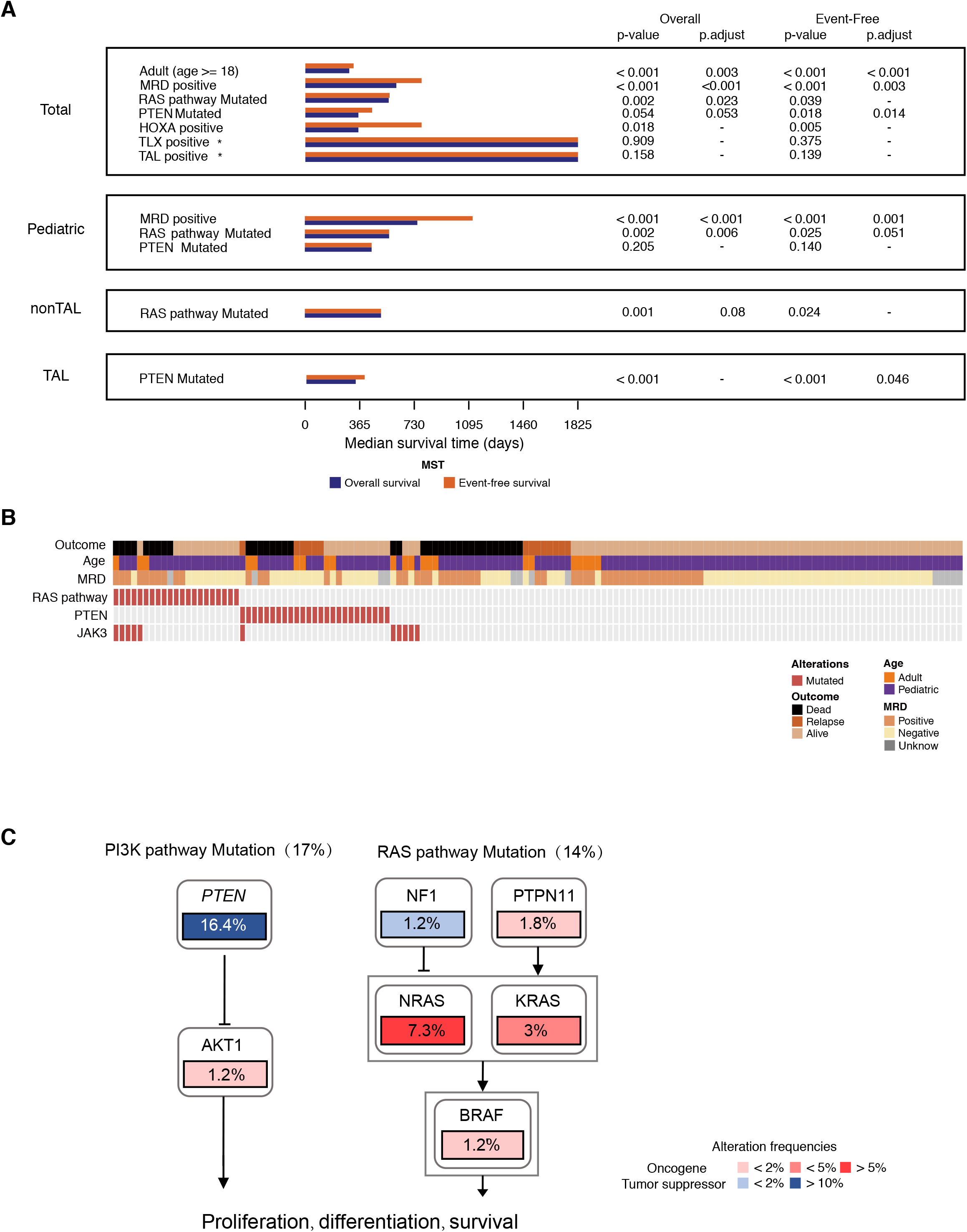
Multi-variable analysis of high-risk factors in T-ALL. (A) Bar graph shows the median overall (blue) and event-free (orange) survival time associated with each clinical or genetic feature; right, p-value and adjusted p-value of overall and event-survival analysis. Adjusted p-values larger than 0.1 were not shown. Symbol * after TLX and TAL represent median survival time longer than 5 years (1825 days). (B) Heatmap showed the correlation of major clinical and genetic risk factors. (C) Mutations identified in this cohort that were associated with the *PI3K* (left) and *RAS* (right) pathways.

In our study, mutations in the *RAS* pathway and *PTEN* represented the most significant genetic risk factors and were mutually exclusive from each other (Figures 1C, 5A-B). *RAS* pathway mutations were enriched in non-TAL subtype with the median OS and EFS time around 1 to 2 years, while *PTEN* mutations were preferentially present in TAL subtype with median OS and EFS time around 1 year (Figure 3F-G and Figure 5A-B). Previous studies have identified *RAS/PTEN* mutations as high-risk factors in adult T-ALL[6]. We found in this study that *RAS/PTEN* mutations were also associated with poor outcomes in pediatric T-ALL, and demonstrated the prognostic value of *RAS* pathway and *PTEN* mutations in nonTAL and TAL subtypes, respectively. The mutual exclusive relationship between *RAS* pathway and *PTEN* mutations could be confirmed by retrospectively analyzing data from previous publications[7-9]. Interestingly, although *RAS* pathway and *PTEN* co-mutations were extremely rare in clinical samples (0 in this and Gianfelici[53] studies, 1/168 in Trinquand study[6]), 5/14 T-ALL cell lines documented in the CCLE datasets were *RAS* pathway and *PTEN* co-mutated[54], which may reflect the selective pressure *in vitro* when these lines were established.

Our study also demonstrates the mutual exclusive relationships of *PTEN* mutations with *JAK3* mutations (Figure 5B), as well as mutations-associated with better prognosis, including those in the *NOTCH/FBXW7* pathways, transcription factor *BCL11b* and *USP7* (Figure 1C and Supplemental Figure 3G-H)[8, 55, 56]. Increased NOTCH activity and *HES1* expression have been reported to associate with improved outcome in pediatric T-ALL[57]. Although we also observed a trend of increased *HES1* expression in patients with *NOTCH/FBXW7* pathway mutations, we did not observe significant survival differences between *HES1* high vs. low or *NOTCH/FBXW7* WT vs. mutant patients in the entire population or in pediatric T-ALLs (data not shown).

Besides these genetic mutations, upregulated *HOXA* family transcription factor expressions were also associated with poor prognosis with the median survival time around 1 - 2 years (Figure 3C, and Figure 5A). Our recent study has identified translocation-mediated neo-loops and *NUP98*-related fusion events as underlying mechanisms for 3D genome alterations associated with dysregulated *HOXA13* expression. Interestingly, patients with *HOXA11-A13* expression, but not other genes in the *HOXA* cluster, have poor outcomes[13]. In our cohort, HOXA subtype was preferentially present in adult T-ALL while TLX and TAL subtypes were preferentially present in pediatric T-ALL (Supplemental Figure 3D), which may explain the longer median survival time in these two subtypes.

Although the spectrum of driver alterations we identified in this Chinese cohort was very similar to those reported in the Western cohort using similar methodologies[8], the mutation rate of *NOTCH1* was relatively low in our cohort. Liu et al found that 74.6% pediatric T-ALL patients harbor the *NOTCH1* mutations[8] while only 52% pediatric T-ALL patients in our cohort carried the *NOTCH1* mutations. Similarly, *NOTCH1* mutation rates in two other Chinese T-ALL studies were also lower than that of the Western cohort when same standard was considered[7, 58]. *NOTCH* pathway mutations are generally associated with better prognosis[6, 11, 53], however, we did not observe this correlation in our population (Supplementary Table 5-6). The lower *NOTCH1* mutation rate in the Chinese T-ALLs may contribute to the lower OS and EFS rates as compared to the Western T-ALLs[8].

Our analysis also identified potential actionable targets associated with the high-risk factors. In our cohort, mutations in the *PTEN* and *RAS* pathways accounted for 16% and 14% of cases, respectively (Figure 5C; Supplemental Figure 4). We also demonstrate that T-ALLs with *RAS* pathway mutations are more sensitive to anti-RAS/MAPK pathway inhibitors *in vitro* and *in vivo* (Figure 4). This, together with previously reported sensitivities of *PTEN* mutated T-ALLs to anti-PI3K targeted therapies[30, 50, 51, 59], suggest that *RAS* pathway and *PTEN* mutations may serve as both prognostic indicators and actionable drug targets for more than 30% T-ALLs. Previous study also suggested that the combination of PI3K inhibitor and MEKi inhibitor was an effective treatment strategy in relapse T-ALLs[60]. If we narrowly define those patients who had died or relapsed within three years as high-risk patients, then 43% of them had potential actionable targets (Supplementary Table 9). Further pre-clinical and clinical investigations are required to test these targets.

## Data Availability

The raw sequence data reported in this paper have been deposited in the Genome Sequence Archive in National Genomics Data Center, China National Center for Bioinformation / Beijing Institute of Genomics, Chinese Academy of Sciences, under accession number HRA000122 that are publicly accessible at https://bigd.big.ac.cn/gsa.

## Acknowledgements

The authors thank Meng Lv, Yingjun Chang, Yan Chang,Yongzhan Zhang for sample collections and Yilin Wu, Ningning Yao, and Liuzhen Zhang for their help with the experiments. The authors thank Cheng Li from Peking University for his suggestion on the bioinformatic analysis. The authors also thank Xuefang Zhang and Yan Liu from the National Center for Protein Sciences Beijing at Peking University and Tsinghua University, respectively, for their assistance. We thank Dr. Xiaotu Ma of St. Jude Children’s Research Hospital for cross-checking our mutation calling analysis. Part of the bioinformatics analysis was performed using the High Performance Computing Platform of the Center for Life Science.This project was supported by the National Natural Science Foundation of China (81602254 for L.Y).This work was also supported by the Peking-Tsinghua Center for Life sciences and Beijing Advanced Innovation Center for Genomics at Peking University to HW. Finally, we thank all the patients who provided their samples for this research.

## Authorship Contributions

Contribution: H.Z., B.D. and H. W. designed the experimental plans. H.Z., Y.Z., W.W., L.Y. and A.Y. performed the experiments. B.D., M.W., J.R., B.C. and Y.L. performed the bioinformatic and statistical analyses; Y.Z., Q.J., and Z.L. collected the clinical samples and data. L.Z., X.Z. and X.H. were in charge of the clinical studies. H.Z., B.D., W.M. and H.W. wrote the manuscript with input from all authors.

## Conflict of interest

The authors declare no competing financial interests.

---

**Figure S1.**
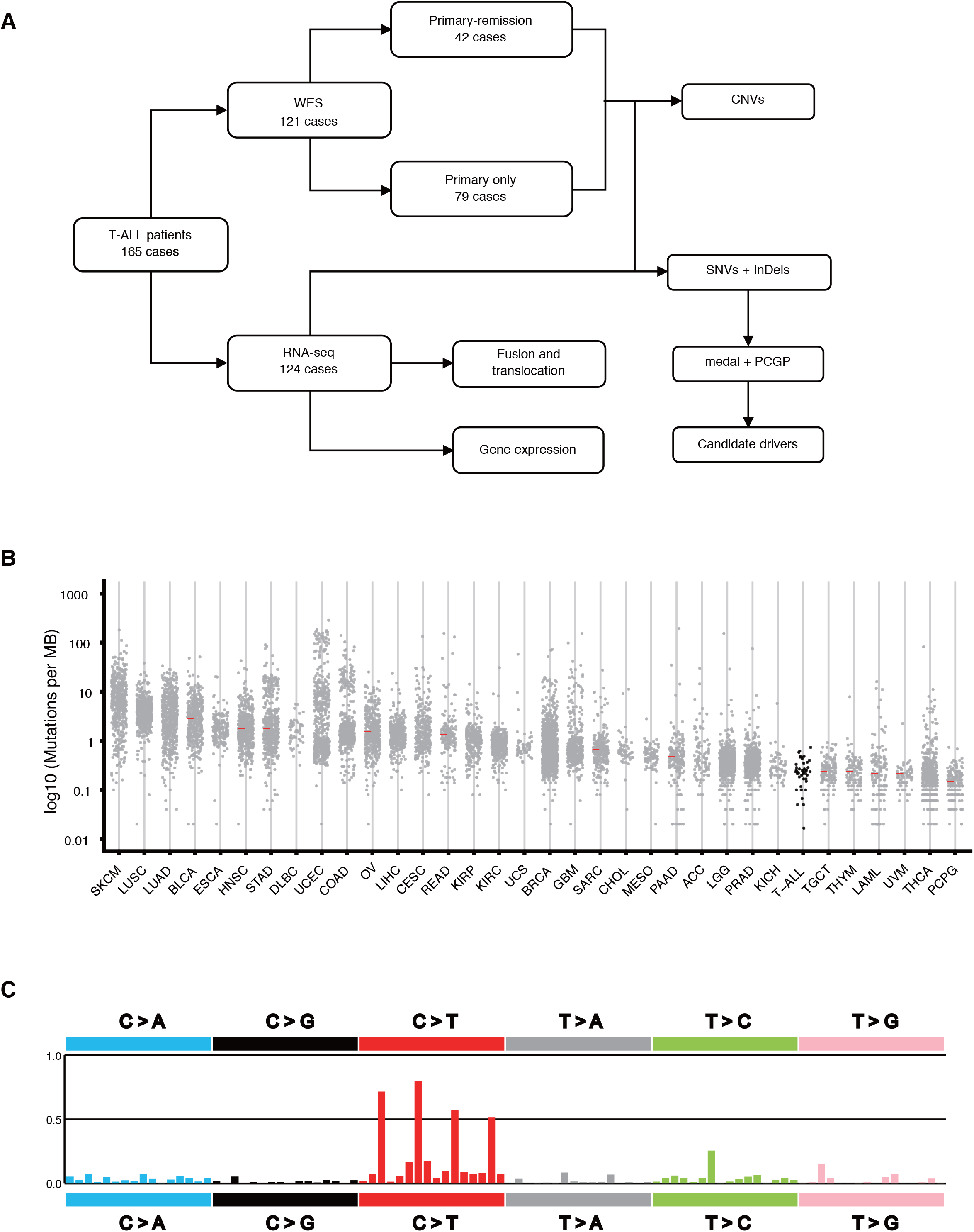
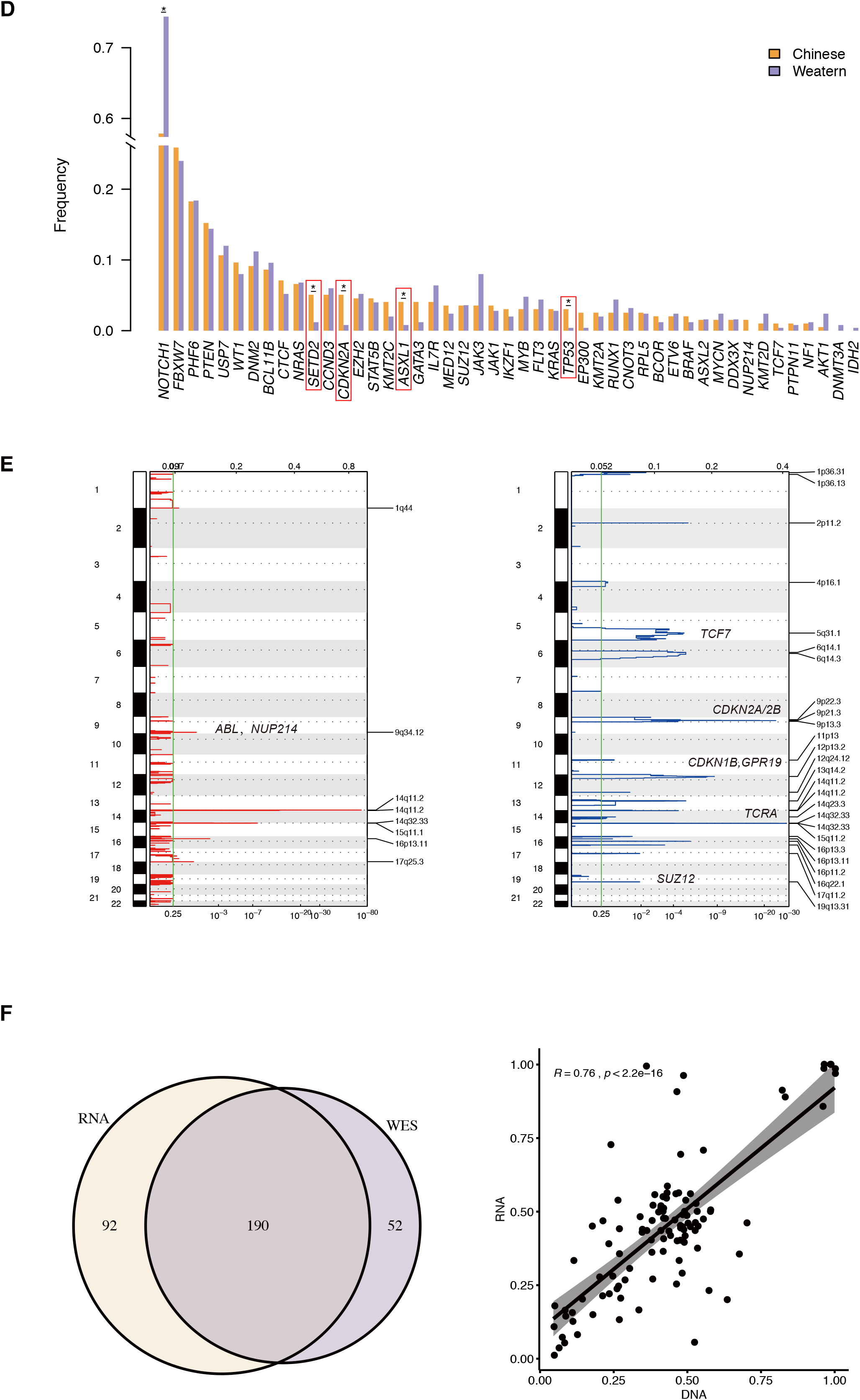
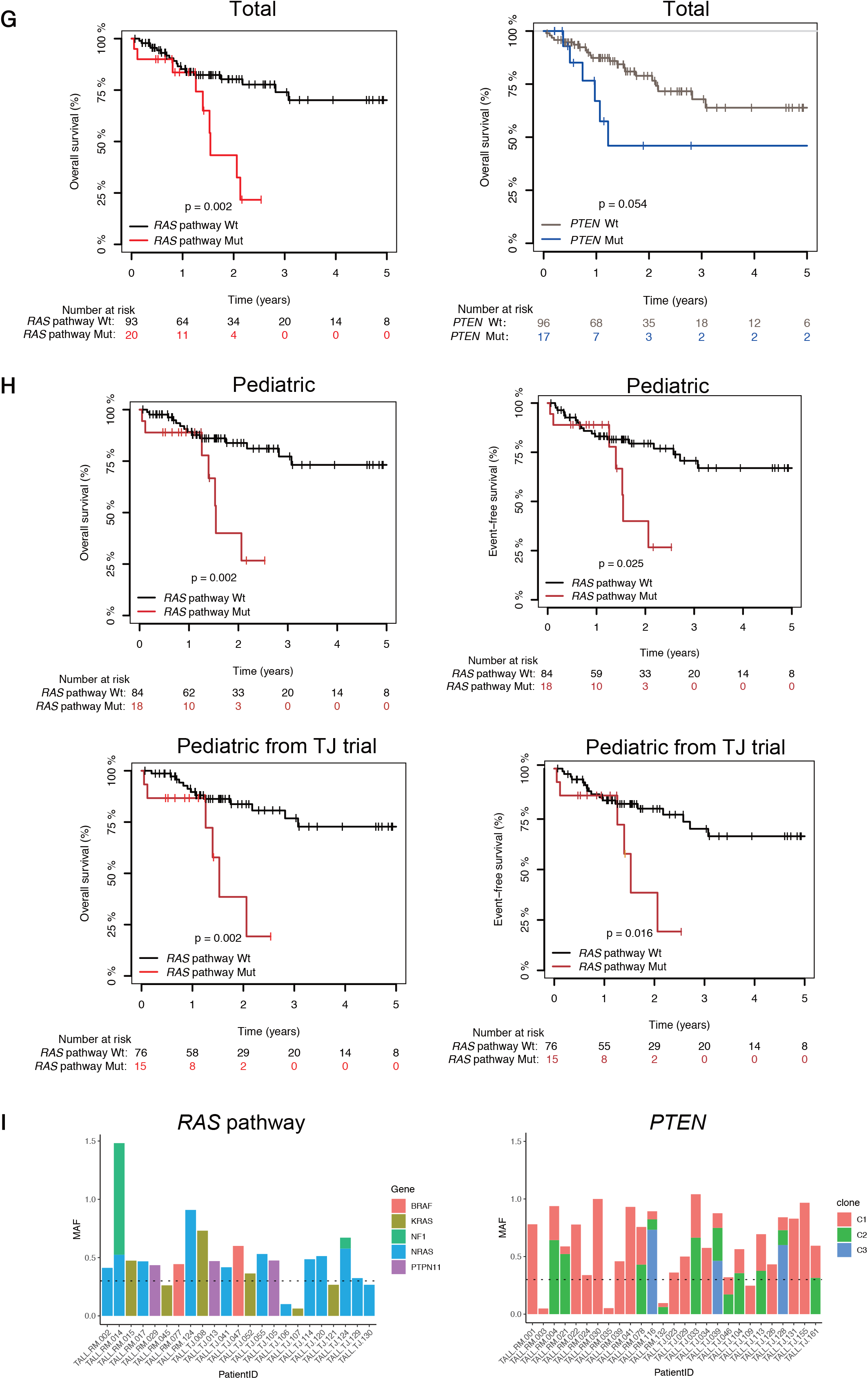

**Figure S2.**
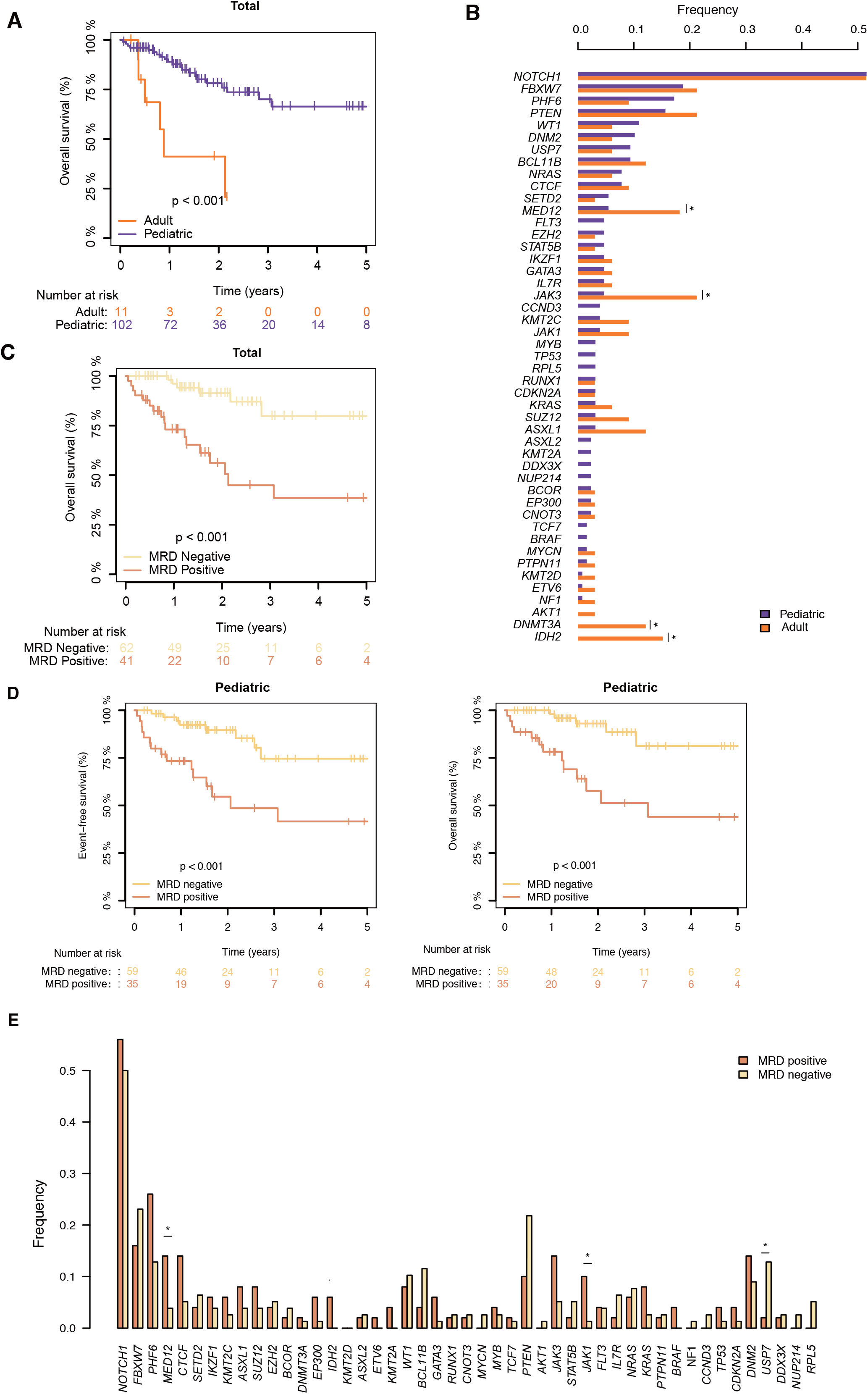
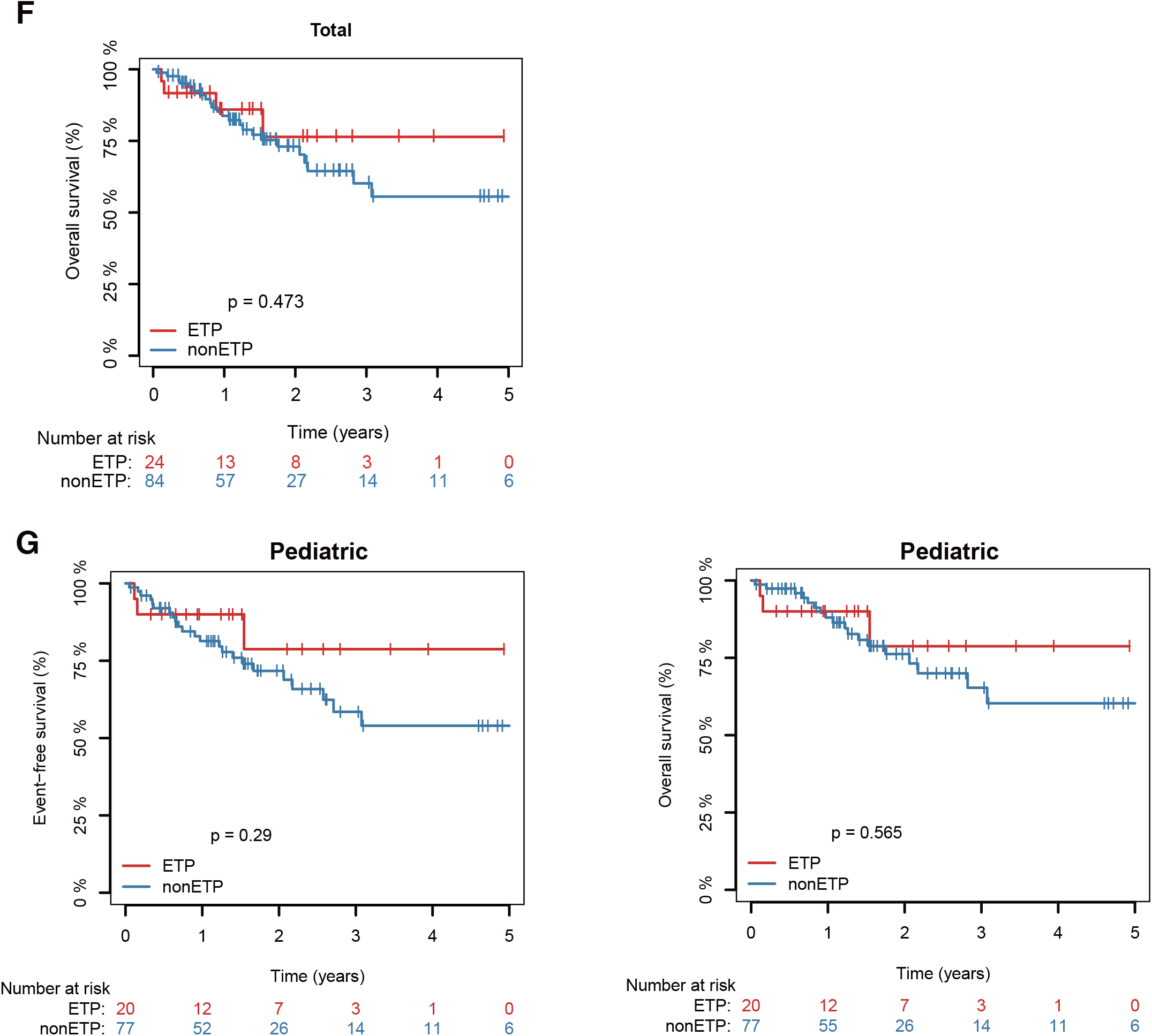

**Figure S3.**
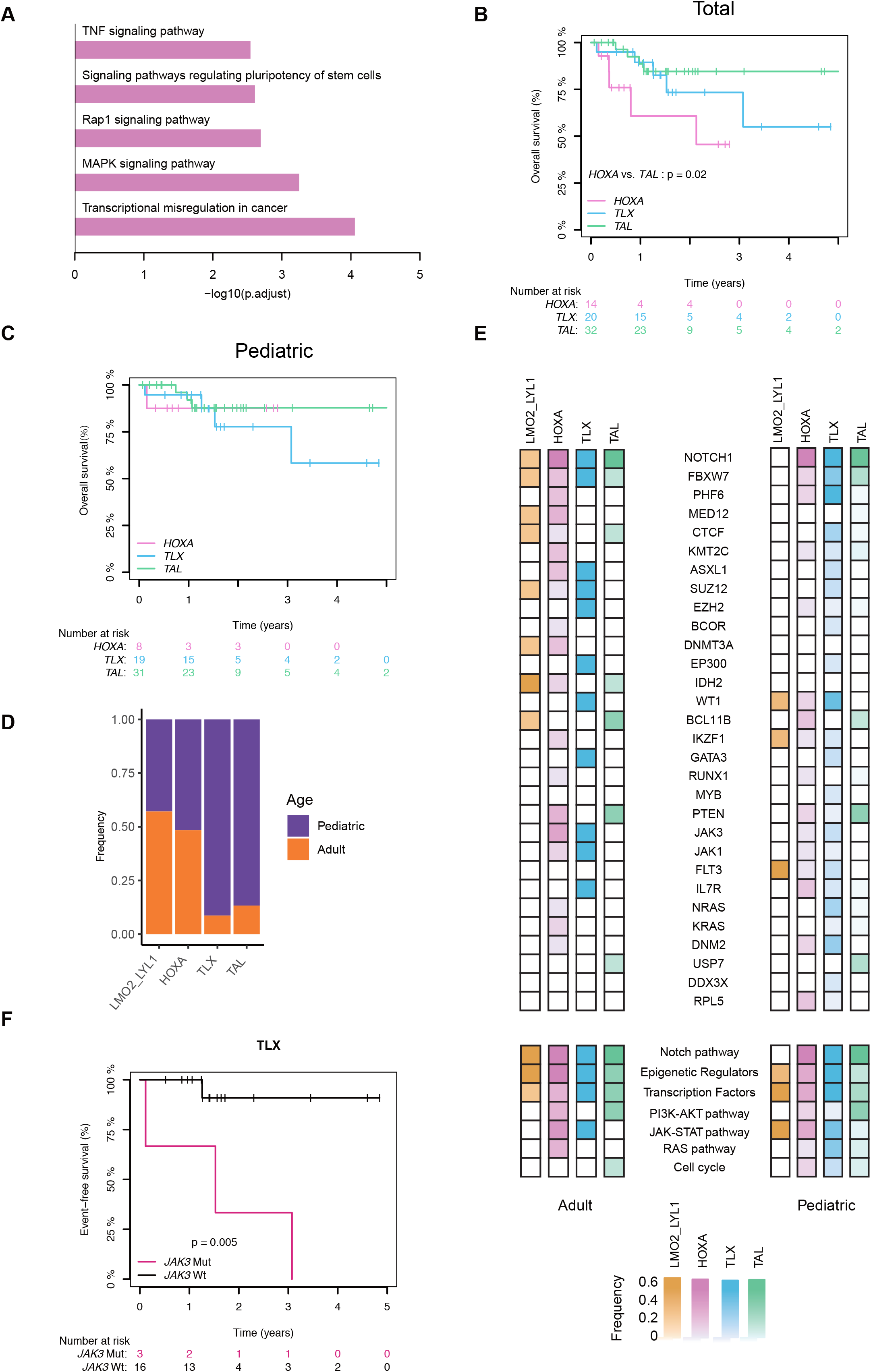
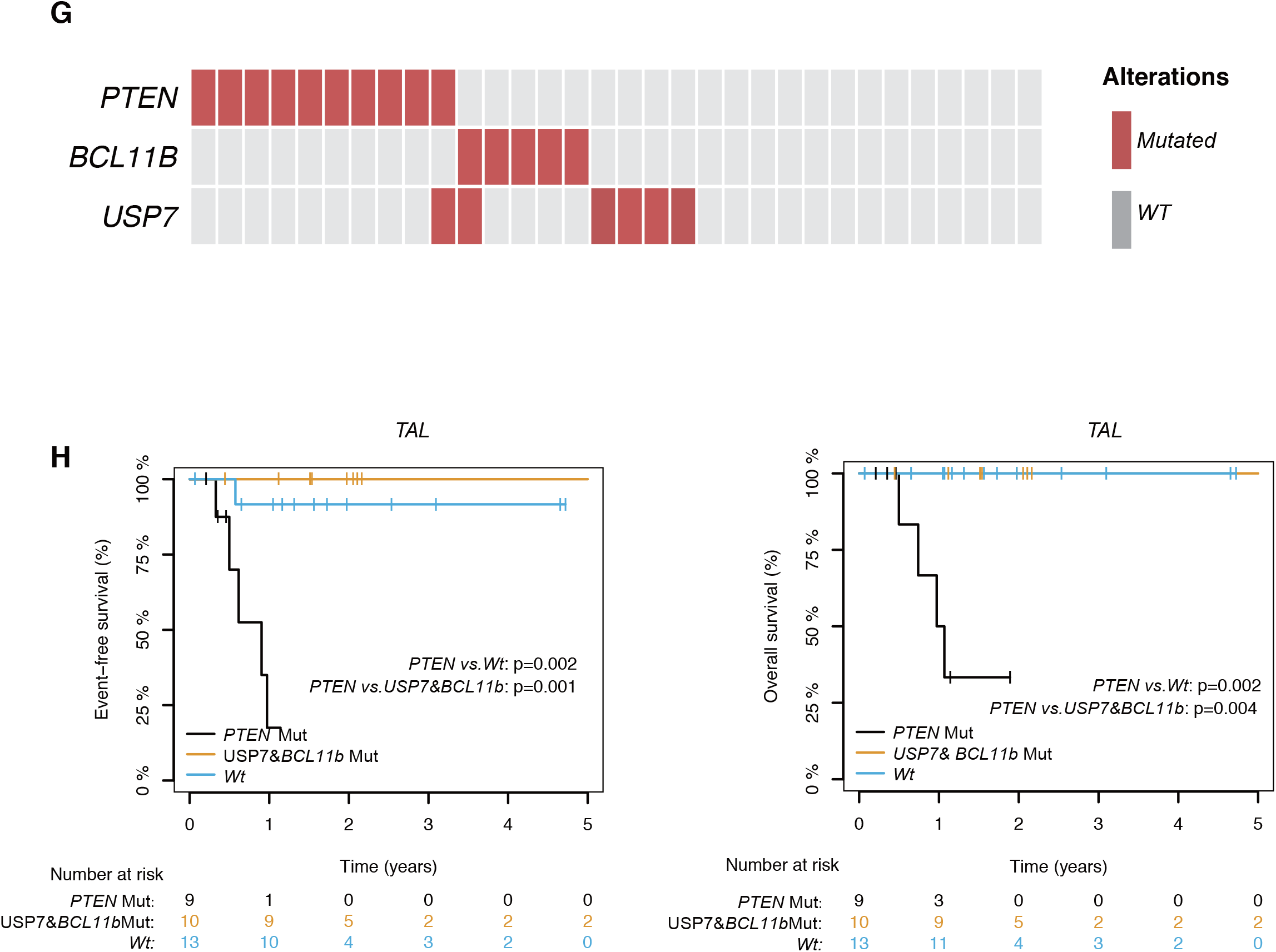

**Figure S4.**
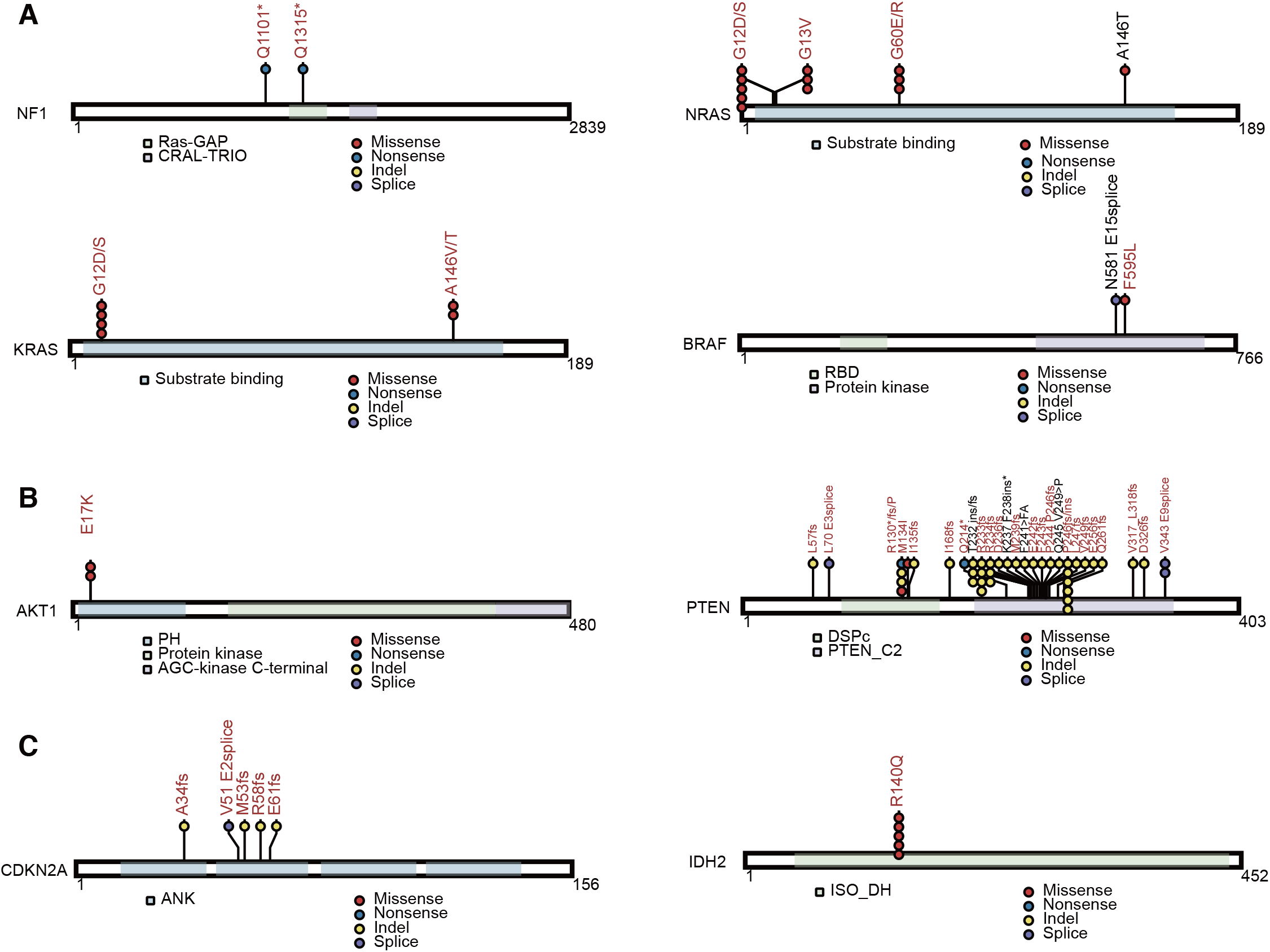

## Notes

### Competing Interest Statement

The authors have declared no competing interest.

### Clinical Trial

NCT00707083; ChiCTR-IPR-14005706;

### Author Declarations

All patients gave written informed consent for treatments and sample collections. The collection of samples and clinical information were approved by the Ethics Committee of Hematology Hospital, Chinese Academy of Medical Sciences and the Ethics Committee of the Peking University People's Hospital.

