## supplementary text for "Integrated Genomic Analyses Identify High-Risk Factors and Actionable Targets in T-Cell Acute Lymphoblastic Leukemia"

**Supplemental Figure Legends**

**Supplemental Figure 1.** (A) Schematic illustration of procedures used for sample analyses. (B) Comparing the overall somatic mutation burden in the Chinese T-ALL cohort and other cancer types. (C) The somatic mutational signature of the Chinese T-ALL cohort. (D) Bar graph shows the gene mutation rates in Chinese (combining data from this study and Chen et al.; orange) and Western (Liu et al.; purple) pediatric T-ALL. (E) Recurrent focal copy-number alterations in T-ALL. The significant (threshold FDR 0.25; green line) focally amplified (red) and deleted (blue) regions are shown. (F) Left, Comparing the driver sites identified by RNA-seq and WES. Right, Correlation of the MAF calculated by RNA-seq and WES. (G) Kaplan-Meier overall survival of T-ALLs with (red) or without (black) *RAS* pathway mutations (left) and with (blue) or without (grey) *PTEN* mutations (right) in the entire cohort. (H) Kaplan-Meier overall (left) and event free (right) survival of T-ALLs with (red) or without (black) *RAS* pathway mutations in the pediatric (upper) and TJ trail patients (lower). (I) Mutated allele frequency of *RAS pathway* (left) and *PTEN* (right) in each patient, different colors within each individual represent different mutation sites. *, $p<0.05$.

**Supplemental Figure 2.** (A) Kaplan-Meier overall survival curve of adult (orange) and pediatric T-ALLs (purple). (B) Bar graph showed the different rates of gene mutation (left) in adult (orange) and pediatric (purple) T-ALLs. (C) Kaplan-Meier overall survival curve of MRD positive (brown) and MRD negative (yellow) T-ALLs. (D) Kaplan-Meier event-free survival curves (left) and overall survival curve (right) of MRD positive (brown) and MRD negative (yellow) in pediatric T-ALLs. (E) Bar graph showed the different rates of gene mutation in MRD positive (brown) and MRD negative (yellow) T-ALLs. (F) Kaplan-Meier overall survival curves of ETP (red) and non-ETP ALLs (blue). (G) Kaplan-Meier event-free survival curves (left) and overall survival curve (right) of ETP (red) and non-ETP ALLs (blue) in pediatric T-ALLs. *, $p<0.05$.

**Supplemental Figure 3.** (A) KEGG analysis of genes enriched in HOXA subtype. (B-C) Kaplan-Meier overall survival of the entire cohort (B) or pediatric T-ALLs (C) of

HOXA subtype (pink), as compared to TLX (blue) and TAL subtypes (green). (D) The proportions of adult and pediatric T-ALLs in LMO2/LYL1, HOXA, TLX and TAL subtypes. (E) Heatmap shows the rates of gene mutation and mutation-associated functional category in LMO2/LYL1 (yellow), HOXA (pink), TLX (blue) and TAL cases (green) in adult (left) and pediatric (right) T-ALLs. (F) Kaplan-Meier event-free survival curve of T-ALLs with (red) or without (black) *JAK3* mutations in TLX subtypes. (G) The mutation status of *PTEN*, *BCL11B* and *USP7* in the TAL subtype patients were enrolled in the survival analysis. (H) Kaplan-Meier event-free survival (left) and overall survival (right) curve of *PTEN* mutation only (blue), *BCL11B* or *USP7* mutation (yellow), and wildtype (black) in TAL subtype.

**Supplemental Figure 4.** Mutation profiles for (A) *NF1*, *NRAS*, *KRAS*, *BRAF;* (B) *AKT1*, *PTEN*; and (C) *CDKN2A* and *IDH2* genes. Potential actionable alterations were marked by red color.

**Supplementary Table Legends**

**Supplementary Table 1.** Clinical information of the patients enrolled in this study.

**Supplementary Table 2.** Mutations detected by WES and RNA-seq data.

**Supplementary Table 3.** Recurrently mutated genes and mutation-associated functional categories.

**Supplementary Table 4.** Significantly mutated genes calculated by MuSiC.

**Supplementary Table 5.** Overall survival analysis on recurrent mutated genes and mutation-associated functional categories in the total population, pediatric population, patients participated in clinical trial, HOXA subtype, TLX subtype and TAL subtype.

**Supplementary Table 6.** Event-free survival analysis on recurrent mutated genes and mutation-associated functional categories in the total population, pediatric population, patients participated in clinical trial, HOXA subtype, TLX subtype and TAL subtype.

**Supplementary Table 7.** Fusion events detected by RNA-seq data.

**Supplementary Table 8.** Expression matrix of the T-ALL patients.

**Supplementary Table 9.** Actionable alterations in the T-ALL patients.

**Supplementary Table 10.** IC50 of the RAS inhibitors of T-ALL cell lines.

**Supplementary Table 11.** Multivariable analyses of overall and event-free survival according to clinical and genetic features and selected variables.
